# Use of ChatGPT in Pediatric Urology and its Relevance in Clinical Practice: Is it useful?

**DOI:** 10.1101/2023.09.11.23295266

**Authors:** Antonio Vitor Nascimento Martinelli Braga, Noel Charlles Nunes, Emanoel Nascimento Santos, Maria Luiza Veiga, Ana Aparecida Nascimento Martinelli Braga, Glicia Estevam de Abreu, Jose Bessa, Luis Henrique Braga, Andrew J Kirsch, Ubirajara Barroso

## Abstract

**Introduction:** Artificial intelligence (AI) can be described as the combination of computer sciences and linguistics, objective building machines capable of performing various tasks that otherwise would need Human Intelligence. One of the many AI based tools that has gained popularity is the Chat-Generative Pre-Trained Transformer (ChatGPT). Due to the popularity and its massive media coverage, incorrect and misleading information provided by ChatGPT will have a profound impact on patient misinformation. Furthermore, it may cause mistreatment and misdiagnosis as ChatGPT can mislead physicians on the decision-making pathway.

**Objective:** Eevaluate and assess the accuracy and reproducibility of ChatGPT answers regarding common pediatric urological diagnoses.

**Methods:** ChatGPT 3.5 version was used. The questions asked for the program involved Primary Megaureter (pMU), Enuresis and Vesicoureteral Reflux (VUR). There were three queries for each topic, adding up to 9 in total. The queries were inserted into ChatGPT twice, and both responses were recorded to examine the reproducibility of ChatGPT’s answers. After that analysis, both questions were combined, forming a single answer. Afterwards, those responses were evaluated qualitatively by a board of three specialists with a deep expertise in the field. A descriptive analysis was performed.

**Results:** ChatGPT demonstrated general knowledge on the researched topics, including the definition, diagnosis, and treatment of Enuresis, VUR and pMU. Regarding Enuresis, the provided definition was partially correct, as the generic response allowed for misinterpretation. As for the definition of VUR, the response was considered appropriate. And for pMU it was partially correct, lacking essential aspects of its definition such as the diameter of the dilatation of the ureter. Unnecessary exams were suggested, for both Enuresis and pMU. Regarding the treatment of the conditions mentioned, it specified treatments to Enuresis that are known to be ineffective, such as bladder training.

**Discussion:** AI has a wide potential to bring several benefits to medical knowledge, improving decision-making and patient education. However, following the reports on the literature, we found a lack of genuine clinical experience and judgment from ChatGPT, performing well in less complex questions, yet with a steep decrease on its performance as the complexity of the queries increase. Therefore, providing wrong answers to crucial topics.

**Conclusion:** ChatGPT responses present a combination of accurate and relevant information, but also incomplete, ambiguous and, occasionally, misleading details, especially regarding the treatment of the investigated diseases. Because of that, it is not recommended to make clinical decisions based exclusively on ChatGPT.

## Introduction

Artificial Intelligence (AI) can be described as the combination of computer sciences and linguistics, with the objective of building machines capable of performing various tasks that otherwise would need Human Intelligence (HI)[1,2]. Therefore, those tasks typically include the ability to understand, learn, adapt, rationalize concrete and abstract concepts, and express complex attributes such as emotions, creativity and problem-solving[2,3].

One of the many AI-based tools that has gained popularity is the Chat-Generative Pre-Trained Transformer (ChatGPT), a chatbot that uses mostly three types of AI. *Analytical AI*, as it identifies, interprets, and communicates in a meaningful way, searching for new insights and data-driven decision-making as it dives through massive quantities of data[1]. *Interactive AI*, as it enables efficient and interactive communication in an automated fashion[1]. In addition, *Textual AI*, as it covers a more natural language processing, therefore having functionalities such as text recognition and answering queries naturaly[1,4].

ChatGPT, was launched by Open AI on November 30, 2022, surpassing 100 million users two months after launch.[4] Due to its Large Language Model (LLM) architecture, analyzing tons of data through Machine Learning (ML) and Deep Learning (DL) using neural networks and predictive models, ChatGPT can answer a quite large array of questions from a massively broad range of topics, including medicine and urology, creating an endless realm of possibilities[1,4,5].

Today, ChatGPT is used thoroughly, as a “Search Engine”, by patients and physicians, seeking adjuvant information and knowledge about their disease [6,7]. However, due to its popularity, and its massive media coverage, incorrect and misleading information provided by ChatGPT will have a profound impact, leading patients, families and physicians to misinformation[6,7]. Furthermore, it may cause misdiagnosis and mistreatment as ChatGPT can lead the physician to the wrong path on the decision-making chart[6–9].

Consequently, it is essential to evaluate and asses its accuracy and consistency regarding pediatric urological queries. The aim of this study is to evaluate and investigate the reliability of ChatGPT 3.5 in terms of concept description, as well as usefulness for decision-making in clinical practice regarding pediatric urology.

## Methods

For this study, the ChatGPT 3.5 version from March 14 was used. The questions asked for the LLM program involved three major pediatric urological conditions, Primary Megaureter (pMU), Enuresis and Vesicoureteral Reflux (VUR).

Regarding the number of questions prompt to ChatGPT, there were 3 queries for each of the topics mentioned above therefore adding up to 9 in total. For Enuresis it was asked: “What is the definition of Enuresis? “How to diagnose Enuresis in a two-page long answer?”; “How to treat Enuresis in a two-page long answer?”. For pMU it was asked: “What is the definition of Primary Megaureter?”; “How to diagnose Primary Megaureter in a two-page long answer?”; “How to treat Primary Megaureter in a two-page long answer?”. In contrast, for VUR the “interview” was as follows: “What is the definition of Vesicoureteral Reflux?”; “What are the indications for endoscopic injection treatment for Vesicoureteral reflux, in a two-page long answer?”; “What are the results for endoscopic injection treatment for Vesicoureteral reflux, in a two-page long answer?”.

Each question was entered as a separate, independent prompt using the “New Chat” function and an anonymous window to minimize bias. The suffix “in a two-page long answer” was used to assure that ChatGPT would provide a similar length answer to every query. The queries were inserted into ChatGPT twice, at the same time, and both responses were recorded to examine the reproducibility of ChatGPT’s answers.

After that analysis of its reproducibility, both questions were combined, forming a single unified answer. Afterwards, those responses were evaluated qualitatively by a board of three specialists, considered thought leaders in the field (AK, LB and UB), each providing a detailed report regarding the accuracy of ChatGPT and its performance.

Descriptive analyses were performed for each urologic condition. The ChatGPT answers were categorized to whether the answers provided impactful or misleading information for both potential patients and healthcare providers. We categorized the ChatGPT’s answer in three types, according to its relevance in clinical practice and patient’s information: 1 – Misleading/Negative impact to care; 2-Helpful to lay person but lacking in substance; and 3-Useful information for patients and treating healthcare providers; and summarized that in Table 1 to objectively elucidate the ChatGPT’s answers.

A table (table A) with ChatGPT’s type of answer was generated. The ChatGPT’s answers are summarized in appendix A.

## Results

The information contained in the ChatGPT responses was analyzed by pediatric urologists who are experts in the field and have extensive clinical experience in the respective pathologies.

### Enuresis

The ChatGPT defines enuresis “as medical term that refers to the involuntary discharge of urine, usually during sleep, in a person who is beyond the age of toilet training. It is commonly known as bedwetting”. However, by definition, enuresis always and not usually occurs during sleep. The usage of the term “usually” in the previous explanation might introduce ambiguity and imprecision.

When ChatGPT is asked about the diagnosis of enuresis, it replies that it can be done through “clinical history, physical examination and complementary tests”. Regarding the clinical history, we obtained the following response: “The doctor will ask about the patient’s symptoms… (Appendix A)”. The specialist highlighted that crucial aspects were omitted during the anamnesis, including the evaluation of airway obstruction, daytime urinary symptoms, and psychological abnormalities, which are essential for making an accurate diagnosis.

The ChatGPT writes that “the physical exam after taking a medical history… (Appendix A)”. The evaluation of the lumbosacral region, which is essential for patients with urinary symptoms, should be highlighted. Furthermore, the possible presence of bladder prolapse or enlarged prostate should not be included.

The complementary exams mentioned, and the respective Chat comments were:

- “Urinalysis: This test analyzes a sample of the patient’s… (Appendix A)”.

We found the following errors and inconsistencies in relation to ultrasonography: The evaluation of the post void residual is not mentioned; there is no indication of performing a urodynamic study for the diagnosis of enuresis, as well as blood tests. Magnetic resonance imaging would only be indicated in very particular situations. In addition, there is no citation of the voiding diary and nocturnal calendar, which are essential tools.

The first paragraph of the treatment response contains inaccurate information regarding the effectiveness of bladder training for treating enuresis. The statement suggesting that bladder training involves encouraging the patient to hold urine for progressively longer periods and gradually increasing the time between urinations is incorrect. The continuation of the answer comes with explanation of specific points of treatment:

- “Understanding the Causes: Before initiating any treatment, it is important to… (Appendix A)”.

Regarding treatment, we found some inconsistency and some missed information. Bladder training does not work for monosymptomatic enuresis. It does not mention the dose, duration of treatment and the main side effect of the medications, that is, hyponatremia for desmopressin and cardiotoxicity for imipramine; nothing is said about the success rate of the alarm. ChatGPT provided satisfactory comments on urotherapy, however, it falls short in terms of medication guidance and incomplete information about the alarm method. The lack of details on medication usage, main side effects, success rate, and duration of use hampers the ability to make informed decisions based on the description of enuresis treatment provided by ChatGPT.

### Vesicoureteral Reflux

We have no comment on the definition of this condition provided by ChatGPT, indicating that no inconsistencies were found in the answer: “Vesicoureteral reflux (VUR) is a medical condition that occurs when urine flows… (Appendix A)”.

On the other hand, the response regarding the indication of endoscopic treatment of VUR presented some incomplete and even wrong information. The first indication cited is for “Vesicoureteral Reflux Grades I-III: Endoscopic injection therapy is indicated for patients with VUR grades I-III… (Appendix A)”. Deflux is FDA approved for grades II-IV and also for duplex ureters that are not mentioned in the text.

For patients with high surgical risk, the ChatGPT says that endoscopic injection therapy may be indicated for patients who are at high risk for surgical complications: “Endoscopic injection therapy is a minimally invasive procedure… (Appendix A)”. This information is false, as the procedures are performed under anesthesia in an operating room. Thus, high surgical risk patients are at the same anesthetic risk.

The chat also mentions the treatment of reflux associated with voiding dysfunction with the following information: “Endoscopic injection treatment may be indicated for children with VUR… (Appendix A)”. This statement is not true, because endoscopic injection does not correct voiding dysfunction per se. It can be used in this situation, but it does not correct the filling and emptying changes, and may result in a lower success rate than children without voiding dysfunction.

When inquired about the outcomes of endoscopic injection for the treatment of reflux, ChatGPT provides a list of points, as outlined below.

Initially, the answer generated talks about preventing recurrent urinary tract infections: “One of the main goals of endoscopic injection treatment for VUR is to prevent recurrent UTIs… (Appendix A)”. It was not considered the experience with more than 5 years of follow-up is 94% of clinical success[10].

Regarding the improvement of kidney function, we found in the answer the explanation that “VUR can cause damage and scarring in the kidneys, or that it can lead to a reduction… (Appendix A)”. Preservation of kidney function is difficult to define, as most grades 1-3 have normal function, so they achieve preserved kidney function.

When high success rates are cited in the excerpt: “Endoscopic injection treatment has been shown to be effective… (Appendix A)”. Again, the long-term data is missing.

The answer also includes information about low complication rates: “Endoscopic injection treatment is a minimally… (Appendix A)”. We are not aware about any ureteral perforation.

It concluded that “endoscopic injection treatment for VUR has been shown to be an effective and minimally invasive treatment… (Appendix A)”. No significantly comment on that.

### Primary Megaureter

Concerning the definition of this condition, ChatGPT has been created in lay-term and imprecise language, avoiding using medical terms such as “ureter”. Furthermore, any definition of Primary Megaureter (pMU) must include the dilatation of the ureter, which is exactly what defines this condition, a discussion that ChatGPT does not mention.

In addition, ChatGPT doesn’t mention any cutoff point of ureteral dilatation, which is greater than 7mm measured behind the bladder on ultrasonographic transverse view. ChatGPT also continuous to be unspecific as it does not classify pMU in refluxing, obstructive or refluxing obstructive megaureter. The AI platform mentions that megaureter is typically diagnosed in childhood, however most cases are diagnosed antenatally.

Regarding the diagnosis of pMU ChatGPT level of accuracy is low, with generic and incorrect statements. ChatGPT doesn’t mention the subtypes that may need a Lasix renal scan (MAG3 or DTPA), a VCUG to rule out VUR or even other tests such as MRU. However, most cases are diagnosed prenatally, therefore physical exam is not required. It also says that the physician needs to palpate the bladder, however, once the problem occurs above the bladder level, bladder is not distended in pMU, showing that ChatGPT gave an incorrect statement.

ChatGPT also says that imaging studies, such as ultrasound is needed; however, its answer is very simplistic, doesn’t mentioning the minimal diameter of the ureter, where it should be measured, the grades of hydronephrosis and other aspects that impacts the medical decision. ChatGPT also recommends doing Voiding cystourethrogram (VCUG), once, according to ChatGPT, it can identify VUR. However, VUR is not a complication of pMU, it is considered one of the 3 types of megaureter, showing a mistake. Renal scan is mentioned by ChatGPT correctly, but it doesn’t mention that its major importance: provide information about the drainage of the urinary system which is essential in cases of pMU. ChatGPT also says that urine culture is necessary, but doesn’t show the indications of the exam, that’s only needed if the child develops a UTI, not being necessary for every patient with pMU. It implied that most patients should have a urine test done as part of the diagnostic work up, which is incorrect. Tests such as renal functional such as glomerular filtration rate (GFR) is also mentioned; however, most children will not need these tests, as the condition is unilateral. The major mistake of ChatGPT is that it doesn’t mention the prenatal diagnosis, once there is no way to suspect that a baby has pMU, and most patients are born asymptomatic.

About the treatment options, ChatGPT says correct sentences, such as, it depends on the severity of the condition and any associated complications, the age and overall health of the patients, and that the goals of treatment are to prevent infections, protect kidney function and relieve symptoms if present. However, it doesn’t mention the watchful waiting on antibiotics, once evidence has shown that there is a role for antibiotic prophylaxis in patients with pMU, reducing the rate of UTI.

Regarding endoscopic surgery, ChatGPT states that it corrects the dilatation of the ureter, which is not correct. It is the opposite once it corrects the narrow distal segment of the ureter by stretching it with a balloon or a stent. ChatGPT also states that laparoscopic surgery results in less pain and faster recovery rates; however, this has not been shown in cases of tapered ureteral reimplantation. No comparative studies have been done in the pediatric population, not been possible to state a sentence like that. It also says that nephrectomy is indicated in some cases that the affected kidney isn’t functioning properly or causing complications; however, it is extremely unlikely to have a kidney completely damaged due to pMU. The dilated ureter provides a buffer sparing the kidney from damage as it occurs in ureteropelvic junction obstruction cases.

## Discussion

According to Evidence Based Medicine medical decisions should be based on the latest medical research evidence to provide the most appropriated treatment and diagnosis plan for the patient[11]. AI has the potential to bring several benefits in medical knowledge, such as improving clinical decision-making and contributing to education, once by making direct questions, ChatGPT gives almost instant answers to medical questions based on high level evidence.

However, there are concerns related to excessive confidence in technology and ethical issues in its use. At this time, ChatGPT lacks genuine clinical experience and judgment, and may provide wrong information. Pediatric Urology is a field of medicine with complex pathologies that doesn’t necessarily have direct answers and unique diagnosis or types of treatment, making the clinician’s experience indispensable. ChatGPT performs well in less complex questions; however, its performance decreases as the complexity of medical decisions increases. It demonstrates knowledge equivalent to a third-year medical student, as shown on Aidan Gilson et al’s study, based on its performance in United States Medical Licensing Examination® responses[12].

Several studies have analyzed the impact of ChatGPT on pediatric medical research. Numerous cardiology and oncology approaches have demonstrated the utility of AI, particularly in identifying and classifying disease phenotypes and improving predictive outcome models by incorporating unstructured data. [13–16]. Using AI to identify inhaler techniques in electronic health records for asthma care, a study suggests it may be possible to eliminate the expensive manual chart review required for guideline-conformant documentation in asthma care by employing a machine learning strategy [13]. However, to the best of our knowledge, no study has evaluated the impact of ChatGPT on medical decision-making in Pediatric Urology. This could lead to a greater efficacy of medical information for the patient, a higher rate of treatment adherence, and a reduction in treatment costs, as well as the secondary effects of incorrectly treating prevalent diseases, such as the ones we analyzed.

In our study, we analyzed the quality of the information provided by ChatGPT in responses on relevant topics in Pediatric Urology. These responses were evaluated by globally recognized experts in their respective fields. ChatGPT demonstrated general knowledge on the researched topics, including the definition, diagnosis, and treatment of three common pathologies in pediatric urology: Enuresis, Vesicoureteral Reflux, and Megaureter. However, ChatGPT provided wrong answers to important topics, such as, the definition of enuresis, which was partially correct. It gave generic responses that did not align with reality. Concerning the evaluation and diagnosis, ChatGPT omitted crucial aspects of anamnesis, which is an essential part of clinical evaluation and defines decision-making.

After analyzing the response regarding enuresis, we identified unnecessary diagnostic exams being suggested, including urodynamic studies and magnetic resonance imaging. Regarding the endoscopic treatment of vesicoureteral reflux, we had a response that could confuse the reader regarding the method’s efficacy. This is due to the lack of citation of results found in relevant published studies on the subject, as well as suggesting outcomes that are not realistic, such as the resolution of urinary symptoms[17–19]. This error in treatment is repeated in the response about enuresis, which mentions ineffective treatments such as bladder training.

Our findings concur with those of Katharina Jeblick et al., who examined the use of ChatGPT in radiology reports. They found both accurate and incorrect statements, which were categorized into four error categories: misinterpretation of medical terms, imprecise language, hallucination, strange language, and grammatical errors[20]. While we did find accurate information, given ChatGPT’s broad background, the language models employed lack specialized medical understanding and knowledge, resulting in imprecise responses that occasionally contain phrases from previous interactions[21,22].

Therefore, ChatGPT is a revolutionary tool that can facilitate public access to information. However, these technologies must be upgraded to enhance the comprehension of medical questions and facilitate clinical medical decision-making by providing more specific answers and fewer generic texts.

## Conclusion

We observed that the ChatGPT responses contain a mosaic of accurate and pertinent information; however, the majority of its responses contain broad, insufficient, and misleading information. In the face of the experts’ feedback and evaluations, it is not recommended to base clinical and therapeutic decisions solely on ChatGPT’s knowledge. It is important to disseminate this information to non-expert professionals and patients, given that ChatGPT has received significant media attention and is widely accessible to the public. In this way, we seek to protect users from harm.

## Data Availability

All data produced in the present study are available upon reasonable request to the authors.

---

**Table A.** Questions to ChatGPT according to pathology and its level of response.

| PATHOLOGY | QUESTION | CHATGPT'S TYPE OF ANSWER |
| --- | --- | --- |
| ENURESIS | What is the definition of Enuresis? | Type 2 |
|  | How to diagnose Enuresis in a two-page long answer? | Type 1 |
|  | How to treat Enuresis in a two-page long answer? | Type 1 |
| VESICoureTERAL REFLUX | What is the definition of Vesicoureteral reflux? | Type 3 |
|  | What are the indications for endoscopic injection treatment for Vesicoureteral reflux in a two-page long answer? | Type 1 |
|  | What are the results for endoscopic injection treatment for Vesicoureteral reflux, in a two-page long answer? | Type 2 |
| PRIMARY MEGAURETER | What is the definition of Primary Megaureter? | Type 2 |
|  | How to diagnose Primary Megaureter in a two-page long answer? | Type 1 |
|  | How to treat Primary Megaureter in a two-page long answer? | Type 1 |

Subtitle:

Type 1 - Misleading/Negative Impact to Care.

Type 2 - Helpful to lay person but lacking in substance.

Type 3 - Useful information for patients and treating healthcare providers.

