## Appendix A for "Use of ChatGPT in Pediatric Urology and its Relevance in Clinical Practice: Is it useful?"

### Appendix A - Questions to ChatGPT and its complete answer.

#### Enuresis

**What is the short definition of Enuresis?**

Enuresis is a medical term that refers to the involuntary discharge of urine, usually during sleep, in a person who is beyond the age of toilet training. It is commonly known as bedwetting.

**How to diagnose Enuresis, in a two page long answer?**

Enuresis is a medical condition characterized by involuntary urination during sleep, typically at night. It is also known as bedwetting, and it affects both children aged five years or older, and adults. Diagnosing enuresis involves a thorough evaluation of the patient's medical history, physical examination, and certain diagnostic tests. In this answer, we will describe the steps involved in diagnosing enuresis in detail.

Step 1: Taking a Medical History The first step in diagnosing enuresis is taking a thorough medical history of the patient. The doctor will ask about the patient's symptoms, including how often they experience bedwetting, how long it has been going on, any recent stressful events and whether there are any triggers or factors that seem to make it worse. The doctor will also ask about any medical conditions the patient has, including any history of urinary tract infections, constipation, or other health issues. Additionally, the doctor will ask about any medications the patient is taking, as some medications can cause enuresis and and family history of enuresis.

Step 2: Physical Examination After taking a medical history, the doctor will perform a physical examination to look for any physical abnormalities or underlying health conditions that may be causing enuresis. The doctor will examine the patient's abdomen, genitals, and lower back to check for any signs of injury, infection, or other medical conditions. The doctor may also perform a neurological examination to check for any nerve damage that could be contributing to the patient's bedwetting. The examination should include a thorough evaluation of the patient's urinary tract, including the bladder and kidneys. The physician should look for signs of inflammation, such as redness, swelling, or tenderness. They should also check for any abnormalities in the genital area, such as a prolapsed bladder or an enlarged prostate.

Step 3: Diagnostic Tests, If the medical history and physical examination do not provide a clear diagnosis, may be necessary to diagnose enuresis accurately., the doctor may order diagnostic tests to further evaluate the patient's condition. The physician may also order a urine culture to identify the specific bacteria causing an infection.These tests may include:

- Urinalysis: This test analyzes a sample of the patient's urine to look for signs of infection, inflammation, or other abnormalities.
- Ultrasound: This test uses sound waves to create images of the patient's urinary tract and can help detect any abnormalities or blockages.
- Urodynamic testing: This test involves measuring the pressure and flow of urine as the patient urinates to assess the function of the bladder and urethra. (Urodynamic studies: to measure the bladder's capacity and function, and to identify any abnormalities in the urinary tract)
- Sleep study: This test involves monitoring the patient's brain waves, breathing, and muscle activity during sleep to evaluate the patient's sleep patterns and to determine if there are any underlying sleep disorders that could be contributing to the bedwetting.
- Blood tests: to check for underlying medical conditions, such as diabetes or kidney disease.
- MRI

Step 4: Referral to a Specialist If the diagnostic tests do not provide a clear diagnosis, the doctor may refer the patient to a specialist, such as a pediatric urologist or a sleep specialist. These specialists can perform additional tests and provide more specialized treatment options to help manage the patient's enuresis.

Treatment:

The treatment of enuresis depends on the underlying cause. In some cases, behavioral modification techniques, such as bladder training, may be effective. This involves encouraging the patient to hold their urine for progressively longer periods, gradually increasing the time between urination.

Medications may be prescribed to treat underlying medical conditions, such as bladder or kidney infections or diabetes. Antidepressants may be used to treat enuresis that is related to psychological factors, such as anxiety or depression.

In some cases, a bedwetting alarm may be recommended. This is a device that sounds an alarm when the patient begins to urinate, waking them up and helping them to learn to recognize the sensation of a full bladder and wake up to use the bathroom.

In conclusion, diagnosing enuresis requires a comprehensive evaluation of the patient's medical history, physical examination, and diagnostic tests. By working with their doctor and undergoing these evaluations, patients can receive an accurate diagnosis and appropriate treatment to help manage their condition. Treatment depends on the underlying cause and may include behavioral modification techniques, medications, or other therapies.

**How to treat Enuresis, in a two page long answer?**

Enuresis, commonly known as bedwetting, is a condition that affects individuals of all ages, but it is most common in children. It is defined as the involuntary discharge of urine during sleep, and it can be a source of embarrassment and shame for those who suffer from it. There are several approaches to treating enuresis, including behavioral therapy, medications, and alarms. In this two-page answer, we will discuss these approaches and provide some tips for managing enuresis.

Understanding the Causes:

Before initiating any treatment, it is important to understand the underlying causes of enuresis. Bedwetting can be caused by a range of factors such as delayed bladder maturation, urinary tract infections, psychological stress, constipation, and sleep disorders. Addressing the underlying cause can often lead to effective treatment of enuresis.

Behavioral Therapy:

Behavioral therapy is a type of treatment that focuses on changing the behavior of the person with enuresis. It involves a combination of techniques, including bladder training, timed voiding, and motivational strategies. Bladder training involves teaching the individual to hold urine for longer periods of time, gradually increasing the time between urination. Timed voiding involves establishing a regular schedule for urination, such as every two hours. Motivational strategies may include rewards for dry nights or positive reinforcement for following the treatment plan.

Medications:

Medications may be prescribed for individuals with enuresis who have not responded to behavioral therapy. The most common medications used are desmopressin and imipramine. Desmopressin is a synthetic hormone that reduces the amount of urine produced by the kidneys, while imipramine is an antidepressant that has been shown to improve bladder control. These medications can be effective, but they can also have side effects, such as headaches, nausea, and dizziness.

Alarms:

Bedwetting alarms are another option for treating enuresis. These alarms work by waking the individual when they begin to urinate during sleep. Over time, the individual learns to recognize the sensation of a full bladder and wakes up before they start to urinate. Bedwetting alarms are most effective when used in combination with behavioral therapy. It is important to note that bedwetting alarms can take several weeks or even months to be effective.

Psychological Support:

For some children, enuresis can be caused by psychological factors such as anxiety or stress. In such cases, psychological support such as therapy or counseling may be helpful in addressing the underlying issues.

Encouragement and Support:

It is important to approach the treatment of enuresis with patience and understanding. Encouraging and supporting the child, rather than punishing or shaming them, can help to build their confidence and reduce any anxiety they may feel about bedwetting.

Tips for Managing Enuresis:

There are several things that can be done to manage enuresis, in addition to the treatment options discussed above. These tips include:

1. Limiting fluids before bedtime: It is important to avoid drinking too much before bedtime, as this can lead to a full bladder and increase the likelihood of bedwetting. This technique can be particularly effective for children who wet the bed at specific times during the night.
2. Encouraging regular urination: Encourage the individual to urinate regularly, even if they do not feel the need to go.
3. Protecting the bedding: Use waterproof bedding or protective covers to prevent damage to the mattress.
4. Talking to a healthcare provider: If enuresis persists despite these strategies, it is important to talk to a healthcare provider. They may recommend additional treatment options or refer the individual to a specialist.
5. Dietary Changes: Making changes to the child's diet can also help to treat enuresis. For example, reducing the child's fluid intake before bedtime, avoiding caffeine and sugary drinks, and ensuring they eat a healthy and balanced diet can all help to reduce the likelihood of bedwetting.

In conclusion, enuresis can be a challenging condition to manage, but there are several treatment options available a holistic approach that takes into account the child's physical, psychological, and emotional needs is important. By understanding the underlying causes, and utilizing a range of treatment techniques such as behavioral therapy, medications, and alarms are all effective approaches to treating enuresis, and there are several strategies that can be used to manage the condition. With the right treatment and support, individuals with enuresis can successfully manage their symptoms and improve their quality of life.

**Vesicoureteral Reflux**

**What is the definition of Vesicoureteral Reflux?**

Vesicoureteral reflux (VUR) is a medical condition that occurs when urine flows backwards from the bladder to the ureters, which are the tubes that carry urine from the kidneys to the bladder. Normally, the valve-like mechanism at the junction of the ureter and bladder prevents urine from flowing back into the ureters as urine flows from the kidneys through the ureters to the bladder, and then out of the body through the urethra. However, in people with VUR, this valve mechanism is weakened or absent, allowing urine to flow back up the ureters and potentially reach the kidneys. This can cause infections, scarring, and damage to the kidneys over time. VUR is a common condition that is often diagnosed in children, but can also occur in adults and can be caused by a congenital defect or other abnormalities in the urinary tract.

**What are the indications for endoscopic injection treatment for Vesicoureteral Reflux, in a two page long answer?**

Endoscopic injection treatment, also known as endoscopic injection therapy, is a minimally invasive procedure used to treat vesicoureteral reflux (VUR). The procedure involves injecting a bulking agent into the wall of the ureter at the point where it enters the bladder, creating a valve-like mechanism to prevent urine from flowing back up into the kidney. Endoscopic injection therapy is often considered as a first-line treatment for VUR, or who have not responded to or are not candidates for other treatments, in children and may also be used in select adult patients. The indications for endoscopic injection treatment for VUR are as follows:

1. VUR Grades I-III: Endoscopic injection therapy is indicated for patients with VUR grades I-III, which represent mild to moderate reflux. These grades of VUR are typically associated with a lower risk of kidney damage and are more likely to resolve spontaneously, especially in young children. Endoscopic

injection therapy is often preferred over surgical intervention for these patients as it is minimally invasive and associated with a lower risk of complications.

2. Recurrent Urinary Tract Infections: Patients with VUR who experience recurrent urinary tract infections (UTIs) are also candidates for endoscopic injection therapy. UTIs can lead to kidney damage and other serious complications, and preventing recurrent UTIs is essential to preserve kidney function. Endoscopic injection therapy can help to reduce the risk of recurrent UTIs by preventing urine from flowing back into the kidney.
3. Conservative Management Failure: Children with VUR are typically treated with antibiotic prophylaxis to prevent urinary tract infections (UTIs) and to reduce the risk of kidney damage. Endoscopic injection therapy may be indicated for patients with VUR who have failed conservative management, such as antibiotic prophylaxis or watchful waiting. Conservative management may not be effective in all patients, and some may require more aggressive intervention to prevent complications and preserve kidney function. Endoscopic injection therapy is an effective treatment option for these patients, as it can help to reduce the risk of recurrent UTIs and prevent further kidney damage.
4. High Surgical Risk: Endoscopic injection therapy may be indicated for patients who are at high risk for surgical complications. Endoscopic injection therapy is a minimally invasive procedure that can be performed under local anesthesia, which makes it a safer alternative to open surgical intervention for patients with certain medical conditions or other risk factors. Additionally, endoscopic injection therapy is associated with a shorter recovery time and fewer complications than open surgical intervention, making it an attractive option for patients who want to avoid more invasive procedures. For example, children with certain medical conditions, such as bleeding disorders or severe allergies, may not be able to undergo traditional open surgery.
5. Reflux Associated with Voiding Dysfunction: Endoscopic injection treatment may be indicated for children with VUR who also have voiding dysfunction, a condition in which the bladder does not empty properly during urination. In these cases, endoscopic injection treatment can help to correct the underlying voiding dysfunction and prevent reflux.

6. Family Preference: In some cases, endoscopic injection treatment may be preferred by families who are looking for a less invasive treatment option for their child's VUR. Endoscopic injection treatment is typically performed on an outpatient basis and involves minimal pain and discomfort, making it a desirable option for some families.

In summary, endoscopic injection therapy is an effective treatment option for patients with VUR who have mild to moderate reflux, recurrent UTIs, failed conservative management, reflux unresponsive to antibiotic prophylaxis, reflux associated with voiding dysfunction, high-risk surgical candidates, and families who prefer a less invasive treatment option. The procedure is minimally invasive and associated with a lower risk of complications than open surgical intervention, making it an attractive option for both children and select adult patients. However, the decision to pursue endoscopic injection therapy should be made on a case-by-case basis and after consultation with a urologist or other medical professional.

**What are the results for endoscopic injection treatment for Vesicoureteral reflux, in a two page long answer?**

Endoscopic injection treatment for vesicoureteral reflux (VUR) is a minimally invasive procedure that involves the injection of a bulking agent into the ureterovesical junction (UVJ) to create a valve-like mechanism and prevent the backward flow of urine from the bladder into the ureters and kidneys. The procedure has been shown to be effective in preventing recurrent urinary tract infections (UTIs) and preserving kidney function in patients with low to moderate grade VUR (grade I to III on the International Reflux Study grading scale).

The results of endoscopic injection treatment for VUR vary depending on a number of factors, including the severity of the VUR, the type of bulking agent used, and the experience of the surgeon performing the procedure. However, several studies have reported positive outcomes following endoscopic injection treatment for VUR, including:

1. Prevention of Recurrent UTIs: One of the primary goals of endoscopic injection treatment for VUR is to prevent recurrent UTIs. Several studies have

reported a significant reduction in UTI recurrence rates following the procedure, with one study reporting a 95% reduction in UTI rates over a median follow-up period of 43 months. For example, a study published in the Journal of Pediatric Urology found that the procedure was successful in preventing UTIs in 87% of patients with grade I to III VUR. Another study published in the Journal of Urology found that endoscopic injection treatment was successful in preventing UTIs in 78% of patients with grade I to III VUR.

2. Improvement in Renal Function: VUR can cause kidney damage and scarring, which can lead to reduced kidney function over time. Endoscopic injection treatment can help to prevent further kidney damage and preserve existing kidney function. Several studies have reported improvements in kidney function following the procedure, with one study reporting a significant increase in the glomerular filtration rate (GFR) in patients with VUR and renal scarring. A study published in the Journal of Urology found that the procedure was successful in preserving kidney function in 96% of patients with grade I to III VUR. Another study published in the Journal of Pediatric Urology found that endoscopic injection treatment was successful in preserving kidney function in 89% of patients with VUR.
3. High Success Rates: Endoscopic injection treatment has been shown to be effective in the majority of cases of low to moderate grade VUR. One study reported a success rate of 93% at 6 months and 85% at 36 months following endoscopic injection treatment with dextranomer/hyaluronic acid (Dx/HA) in patients with VUR.
4. Low Complication Rates: Endoscopic injection treatment is a minimally invasive procedure that is associated with low complication rates. The most common complications are mild and transient, such as fever, urinary tract irritation, and pain. Serious complications are rare, but can include ureteral obstruction, ureteral perforation, and migration of the bulking agent. The risk of complications is generally low, with reported rates of less than 5%.

5. Duration of Effectiveness: The duration of effectiveness of endoscopic injection treatment for VUR varies depending on the type of bulking agent used. The most commonly used bulking agents are dextranomer/hyaluronic acid (Dx/HA) and polydimethylsiloxane (PDMS). Studies have shown that

Dx/HA injection treatment has a success rate of 60-80% at 5 years post-procedure, while PDMS injection treatment has a success rate of 77-89% at 5 years post-procedure.

6. Patient Satisfaction: Overall, endoscopic injection treatment for VUR has been shown to be well-received by patients. A study published in the Journal of Pediatric Urology found that 94% of patients and their families were satisfied with the procedure and would recommend it to others.

It is important to note that the long-term outcomes of endoscopic injection treatment for VUR are still being studied, and the durability of the treatment may vary depending on the type of bulking agent used and other factors. Additionally, endoscopic injection treatment may not be appropriate for all cases of VUR, and more aggressive treatment options may be necessary in cases of high grade VUR or significant kidney damage.

In conclusion, endoscopic injection treatment for VUR has been shown to be an effective and minimally invasive treatment option for patients with low to moderate grade VUR. The procedure can prevent recurrent UTIs, preserve kidney function, and has a high success rate with low complication rates. However, it is important to consider the individual patient's needs and the severity of their condition when determining the most appropriate treatment approach.

**Primary Megaureter**

**What is the definition of Primary Megaureter?**

Primary megaureter, also known as congenital or idiopathic megaureter, is a medical condition characterized by the abnormal enlargement or dilation of the ureter, which is the tube that carries urine from the kidney to the bladder. In primary megaureter, the ureter is enlarged at birth or shortly thereafter, usually due to a congenital abnormality in the way the ureter develops. This can cause problems with the flow of urine and may increase the risk of urinary tract infections and kidney damage if left untreated. This dilation is not caused by any other underlying medical condition, such as a blockage or obstruction, and is instead thought to be due to a developmental abnormality in the muscles of the ureter. Primary megaureter is typically diagnosed in childhood and can affect one or both ureters. In most cases, it does not cause any symptoms, but in some cases, it can lead to urinary tract infections, kidney damage, or other complications. Treatment for primary megaureter may involve monitoring the condition, antibiotics to treat infections, or surgical intervention to correct the dilation of the ureter.

**How to diagnose Primary Megaureter, in a two page long answer?**

The diagnosis of primary megaureter typically involves a combination of medical history, physical examination, diagnostic tests and procedures that are used to evaluate the function and structure of the urinary system. These tests are used to identify the presence of the condition, assess the extent of the dilation, and determine any potential complications that may arise.

Medical History: The first step in diagnosing primary megaureter is to take a detailed medical history of the patient. This includes asking questions about the patient's symptoms, such as frequent urinary tract infections, difficulty urinating, or abdominal pain. The doctor will also ask about any family history of urinary tract disorders or other medical conditions.

Physical Examination: After taking a medical history, the doctor will perform a physical examination of the patient. This may involve palpating the abdomen to check for any abnormalities, such as a distended bladder. The doctor may also perform a rectal exam in boys to check for any abnormalities in the prostate gland.

Diagnostic Tests: Once the medical history and physical examination are complete, the doctor may order diagnostic tests to confirm the diagnosis of primary megaureter. These tests may include:

1. Medical history and physical examination: The first step in the diagnosis of primary megaureter involves taking a detailed medical history and performing a physical examination. The doctor will ask about the patient's symptoms, including any urinary tract infections, as well as any family history of kidney or urinary tract

problems. During the physical examination, the doctor will palpate the patient's abdomen to check for any swelling or tenderness.

2. Imaging studies: Various imaging studies are used to evaluate the structure and function of the urinary system, including the ureter. These tests include:

- Ultrasound: An ultrasound is a non-invasive test that uses sound waves to create images of the internal organs. It is often the first test used to evaluate the urinary system and can identify the presence of dilation or other abnormalities in the ureter.

- Voiding cystourethrogram (VCUG): A VCUG is a test that involves injecting a contrast dye into the bladder and then taking X-ray images as the patient urinates. This test can identify any reflux (backward flow) of urine from the bladder into the ureter, which can be a complication of primary megaureter.

- Magnetic resonance imaging (MRI): An MRI uses a powerful magnet and radio waves to create detailed images of the internal organs. It can provide more detailed information about the structure and function of the urinary system and is often used when other imaging studies are inconclusive.
- Renal scan: A renal scan uses a radioactive tracer to visualize the blood flow and function of the kidneys. This test can help to identify any abnormalities in kidney function that may be associated with primary megaureter.

3. Urine tests: Urine tests are used to evaluate the function of the kidneys and urinary tract. These tests include:

- Urinalysis: A urinalysis is a test that involves analyzing a sample of urine for the presence of bacteria, blood, or other abnormalities.

- Urine culture: A urine culture involves growing any bacteria present in a urine sample to determine the type and extent of an infection.

4. Renal function tests: Renal function tests are used to evaluate how well the kidneys are functioning. These tests include:

- Blood tests: Blood tests are used to evaluate the levels of various substances in the blood, including creatinine and blood urea nitrogen (BUN). Elevated levels of these substances can indicate kidney damage.

- Glomerular filtration rate (GFR): The GFR is a test that measures how well the kidneys are filtering waste products from the blood. A decreased GFR can indicate kidney damage.

In conclusion, the diagnosis of primary megaureter typically involves a combination of medical history, physical examination, and diagnostic tests such as ultrasound, VCUG, renal scan, and MRI. Treatment options depend on the severity of the condition and may include antibiotics or surgery to correct the dilation of the ureter. It is important to seek medical attention if you suspect that you or your child may have primary megaureter, as early diagnosis and treatment can help to prevent complications and improve outcomes.

**How to treat Primary Megaureter, in a two page long answer?**

The treatment options for primary megaureter depend on the severity of the condition and any associated complications, as well as the age and overall health of the patient. The goals of treatment for primary megaureter are to prevent infections, protect kidney function, and relieve symptoms, if present.

The following are some of the treatment options for primary megaureter:

1. Watchful waiting: In many cases, primary megaureter does not cause any symptoms or complications, and may not require any treatment. Instead, the patient may be monitored closely over time to ensure that the condition does not worsen or lead to any complications.
2. Antibiotics: If the patient experiences urinary tract infections as a result of primary megaureter, antibiotics may be prescribed to treat the infection and prevent further complications. In some cases, long-term prophylactic antibiotics may be recommended to prevent recurrent infections.
3. Surgery: Surgical intervention may be necessary to correct the dilation of the ureter and prevent further complications. The type of surgery will depend on the extent of the dilation and any associated complications, and may include:

- Ureteral reimplantation: This surgical procedure involves removing the dilated portion of the ureter and reattaching it to the bladder. It is often recommended for patients with severe dilation or reflux.
- Endoscopic surgery: Endoscopic surgery involves using a thin, flexible tube with a camera and surgical instruments to correct the dilation of the ureter. This procedure is less invasive than ureteral reimplantation, but may not be suitable for all patients.
- Laparoscopic surgery: Laparoscopic surgery involves the use of small incisions in the abdomen to access the ureter and make repairs. This is a minimally invasive option that can result in less pain and faster recovery times.

4. Observation and repeat imaging: In some cases, primary megaureter may resolve on its own without the need for intervention. In these cases, the patient may be observed closely with repeat imaging studies to monitor the progression of the condition over time.
5. Nephrectomy: In rare cases, the affected kidney may need to be removed if it is not functioning properly or is causing complications.
6. Follow-up care: After treatment for primary megaureter, patients will require follow-up care to monitor the condition and ensure that there are no further complications. This may involve regular check-ups with a urologist, imaging studies, and urine tests

In addition to these treatment options, it is important for patients with primary megaureter to maintain good urinary hygiene and follow any additional recommendations from their healthcare provider to prevent infections and protect kidney function. This may include drinking plenty of fluids, urinating regularly, and avoiding irritants to the urinary tract.
